# “I feel like my body is broken”: Exploring the experiences of people living with long COVID

**DOI:** 10.1101/2022.01.20.22269617

**Authors:** Amanda Wurz, S. Nicole Culos-Reed, Kelli Franklin, Jessica DeMars, James G. Wrightson, Rosie Twomey

**Affiliations:** School of Kinesiology, University of the Fraser Valley, Chilliwack, BC, Canada; Faculty of Kinesiology, University of Calgary, Calgary, AB, Canada; Department of Oncology, Cumming School of Medicine, Calgary, AB, Canada; Department of Psychosocial Resources, Tom Baker Cancer Centre, Alberta Health Services, Calgary, AB, Canada; Patient partner; Breathe Well Physio, Calgary, AB, Canada; Department of Clinical Neurosciences, University of Calgary and Alberta Children’s Hospital, Calgary, AB, Canada

**Author notes:** **Corresponding author:** Rosie Twomey, Ph.D.

**Keywords:** Post-acute COVID-19 syndrome, Post-COVID-19 condition Post-viral fatigue, COVID-19

## Abstract

**Background:** Long COVID, an illness affecting a subset of individuals after COVID-19, is distressing and poorly understood. Exploring the experiences of people with long COVID could help inform current conceptualizations of the illness, guide supportive care strategies, and validate patients’ perspectives on the condition. Thus, the objective of this study was to better understand and explore individuals’ experiences with long COVID and commonly reported symptoms, using qualitative data collected from open-ended survey responses.

**Methods:** Data were collected from adults living with long COVID following a confirmed or suspected SARS-CoV-2 infection who participated in a larger observational, online survey. Within the larger survey, participants had the option of answering seven open-ended items. Data from the open-ended items were analyzed following guidelines for reflective thematic analysis.

**Results:** From the 213 who were included in the online survey, 169 participants who primarily self-identified as women (88.2%), aged 40-49 (33.1%), and who had been experiencing long COVID symptoms for ≥ 6 months (58.6%) responded to the open-ended questions. Four overlapping and interconnected themes were identified: (1) *My long COVID symptoms are numerous, hard to describe, and debilitating*, (2) *All aspects of my day-to-day functioning have been impacted*, (3) *I can no longer be physically active*, and (4) *I keep asking for help, but no one is listening, and very little is working*.

**Conclusion:** Findings highlight the complex nature of long COVID and show the ways in which individuals affected by the illness are negatively impacted. Participants recounted struggling and altering their daily activities while managing relapsing-remitting symptoms, an uncertain prognosis, lost pre-COVID identities, and a healthcare system (that does not always offer guidance nor take them seriously). More support and recognition for the condition are needed to help this cohort navigate the process of adapting to long COVID.

## Introduction

The acute impact of SARS-CoV-2 (the virus that causes COVID-19) infection varies widely, with some individuals experiencing no symptoms and others experiencing adverse effects that vary from mild to critical severity [1,2]. Diverse responses to COVID-19 persist beyond the initial presentation, with a subset of the population experiencing complicated, disconcerting, prolonged illness [3], widely known as long COVID [4] (also referred to as post-acute sequelae of SARS-CoV-2 infection). Though much remains unknown about long COVID, it is characterized by multiorgan impairments that span respiratory, cardiovascular, neurological, dermatological, and gastrointestinal systems [5]. Commonly reported symptoms include fatigue, shortness of breath, dry cough, cognitive impairment, headache, heart palpitations, chest tightness, and dizziness [6–8]. Symptoms can co-occur, vary in severity, and be cyclical or episodic in nature [9–11]. Initially, the prevalence and seriousness of chronic symptoms after COVID-19 were underrecognized, contested, or dismissed [12,13], and much of the early research on long COVID was led by patients [14]. Researchers and healthcare providers worldwide now recognize the significant burden associated with long COVID [15,16].

Nevertheless, there is still a relatively poor understanding of the lived experience of long COVID. For example, much of the evidence describing long COVID has come from individuals who were hospitalized for COVID-19 [17–21] or excluded those without a laboratory-confirmed COVID-19 infection, despite numerous reasons why this may not have been accessible [22]. Further, the closed-ended questionnaires used within surveys [14,23] may have failed to collect important aspects of the patient voice. Indeed, despite a highly engaged patient population whose activism collectively made long COVID’s complex symptomatology visible [4,14], few studies have included the patient perspective through qualitative approaches. Of the studies that have used qualitative approaches [24–27], the patient experience has been explored via interview and focus group methodologies with individuals affected by long COVID residing in the United Kingdom. In these studies, patients offer detailed accounts of navigating skepticism from the healthcare system and their social networks, the challenges of managing inconspicuous symptoms, the inability to perform physical activity or activities of daily living, and the importance of seeking refuge and information from similar others [24–27]. However, no studies have used open-ended survey items as a lower burden approach to gathering in-depth information, nor captured experiences of individuals with long COVID residing outside of the United Kingdom. Although interviews and focus groups can enable researchers to probe and further explore patients’ lived experiences, in the case of long COVID, such emotionally, socially, and time-consuming methods could preclude those with the most severe symptoms from participating.

Therefore, using data from a larger online survey conducted in 2021, the specific objective of this sub-study was to better understand and explore individuals’ experiences with long COVID and commonly reported symptoms using qualitative data collected from open-ended survey items.

## Methods

The qualitative data collected, analyzed, and reported herein were collected as part of a larger observational study using an online survey [8], which was approved by the University of Calgary Conjoint Health Research Ethics Board (REB21-0159). Initially, the qualitative data were collected to enable participants to qualify or elaborate on their closed-ended responses, with planned content analysis if data were sufficient. However, upon exporting the qualitative data, it became apparent that rich and important insights were offered and that in-depth reflexive thematic analysis [28] of this survey data [29] was warranted. A pragmatic approach [30] and constructivist paradigm [31] was adopted, wherein participants’ voices were centred and reality was viewed as varied and socially created.

### Participants

Individuals were eligible if they self-identified as (1) an adult aged ≥ 18 years; (2) currently experiencing long-term symptoms due to COVID-19 for at least four weeks since the acute illness or positive COVID-19 test, with symptoms not pre-dating the acute illness; and, (3) having tested positive for COVID-19, or with probable infection (based on an illness mimicking the acute phase of COVID-19, having close contact with a confirmed case, or being linked with an outbreak), in line with the clinical definition for long COVID (i.e., post-COVID-19 condition) [32].

### Procedures

Following Research Ethics Board approval, individuals were recruited internationally to participate in the larger online survey study through clinical networks, advertisements placed on social media sites, and snowball sampling. Advertisements for the study included a link that took potential participants to a ‘study objectives’ page. Here individuals could learn about the study and review the eligibility criteria before being directed to the informed consent page. After informed consent was obtained, participants gained access to the secure online survey housed on Qualtrics.

### Measures

The larger online survey consisted of a socio-demographic and medical questionnaire, five closed-ended questionnaires assessing fatigue, post-exertional malaise, health-related quality of life, breathing discomfort, and physical activity (described and presented elsewhere [8]), and seven open-ended items. The seven open-ended items, which are the focus of this manuscript, were presented after each of the five closed-ended questionnaires, or block of questions, via stems such as “*Please use this space for any other comments about your experience with symptoms that continued or developed after acute COVID-19, or the support you have received for long COVID. This is optional, please skip this question if you have no other comments*.” The survey took approximately 30 minutes to complete (29.2 ± 17.1 minutes), and responses to open-ended questions ranged from 5-290 words.

### Data Analysis

Descriptive statistics were computed for socio-demographic and medical data using Jamovi [33]. Open-ended qualitative data were de-identified, transferred to an Excel spreadsheet, and uploaded to be managed in NVivo (QSR International; 12.6.1). Following guidelines for reflexive thematic analysis [28], two authors (AW, RT) independently familiarized themselves with the data by reading the responses several times. Next, data were coded inductively by a single author (AW) who identified salient features within responses to generate initial codes. Salient features were determined based on their relevance to the topic, this sub-study’s objectives, and the author’s judgment. Similar codes were then grouped together into subthemes and main themes that summarized the raw data and conveyed the salient features. At this point, two authors (AW, RT) convened to review the subthemes and main themes and to challenge one another’s interpretations of the data and explore alternative perspectives. The authors also reviewed the themes to check for internal homogeneity (i.e., data within themes fit together meaningfully) and external heterogeneity (i.e., clear distinctions between themes). A theme table including representative quotes was generated, iterated upon, and circulated to a patient partner and the third author (KF) to critically review. Following this, the three authors (AW, KF, RT) met to further discuss and challenge one another’s understanding of and experience with the data (see Supplementary File 1 for the authors’ reflexivity statement). The theme table was then finalized and sent to all authors to review.

Several strategies were employed to ensure sensitivity to the context, commitment and rigour, transparency and coherence, and impact and importance [34]. For example, the authors sought to ask and answer an important, practical, and timely research question and recruited a sample who could provide firsthand accounts. As well, the stages of the research process have been concisely presented above to elucidate the iterative and reflexive nature of the data collection and analysis process and to enhance transparency. Finally, representative quotes are included herein, and a table was generated so that readers may review and critically reflect upon the authors’ interpretations.

### Patient Involvement

Within the larger observational study, a patient partner was involved (March 2021 onward) to contribute to the study design and interpretation of results. For this sub-study, the same patient partner contributed to design and data analysis and interpretation. As described above, the patient partner and author (KF) critically reviewed and reflected on the theme table and was actively engaged in iterative discussions, aiding the interpretation of findings. The patient partner also contributed to three team meetings to discuss the research objectives, review subthemes and themes, and comment on (and approve of) the final version of the manuscript. These meetings lasted 60 to 75 minutes.

## Results

Of the 213 participants who were included in the larger study, 169 (79%) provided responses to the open-ended items and were included herein. Of note, most (n=105; 62%) provided responses to three or more of the open-ended items. The remaining (n=64; 38%) responded to one or two open-ended items.

### Participants

The socio-demographic and medical characteristics of participants in this sub-study are presented in Table 1. Most participants identified as women (*n*=149; 88.2%), and were aged 30-39 (*n*=36; 21.3%), 40-49 (*n*=56; 33.1%), or 50-59 (*n*=39; 23.1%). The majority identified as White (*n*=158; 93.5%) and from Canada (*n*=63; 37.3%), the United Kingdom (*n*=67; 39.6%), or the United States of America (*n*=26; 15.4%). Most participants (*n*=99; 58.6%) described managing long COVID symptoms for more than 10 months.

**Table 1.**
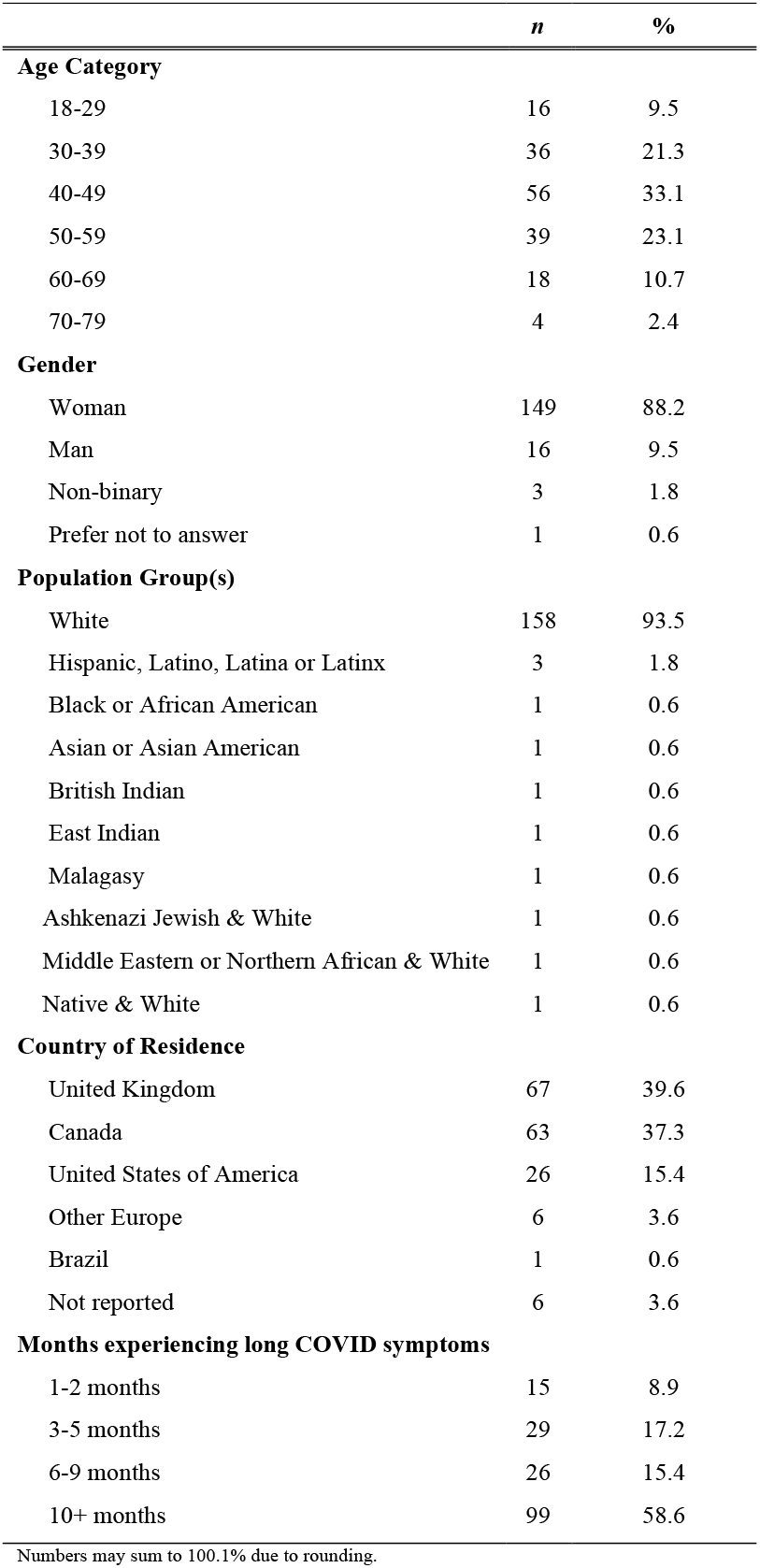
Socio-demographic and medical characteristics of participants.

### Main Results

Participants’ insights into their long COVID experience and commonly reported symptoms are summarized across four themes and eight subthemes, which are described below with illustrative quotes. Table 2 contains additional quotations associated with each theme and subtheme. To respect participant confidentiality, the larger online survey was anonymous, and any potentially identifying information was redacted. As such, no participant numbers nor pseudonyms are used herein. To enhance readability, spelling and grammar were edited, and where necessary, additional descriptive information was added, which is presented within square brackets (i.e., […]).

**Table 2.**
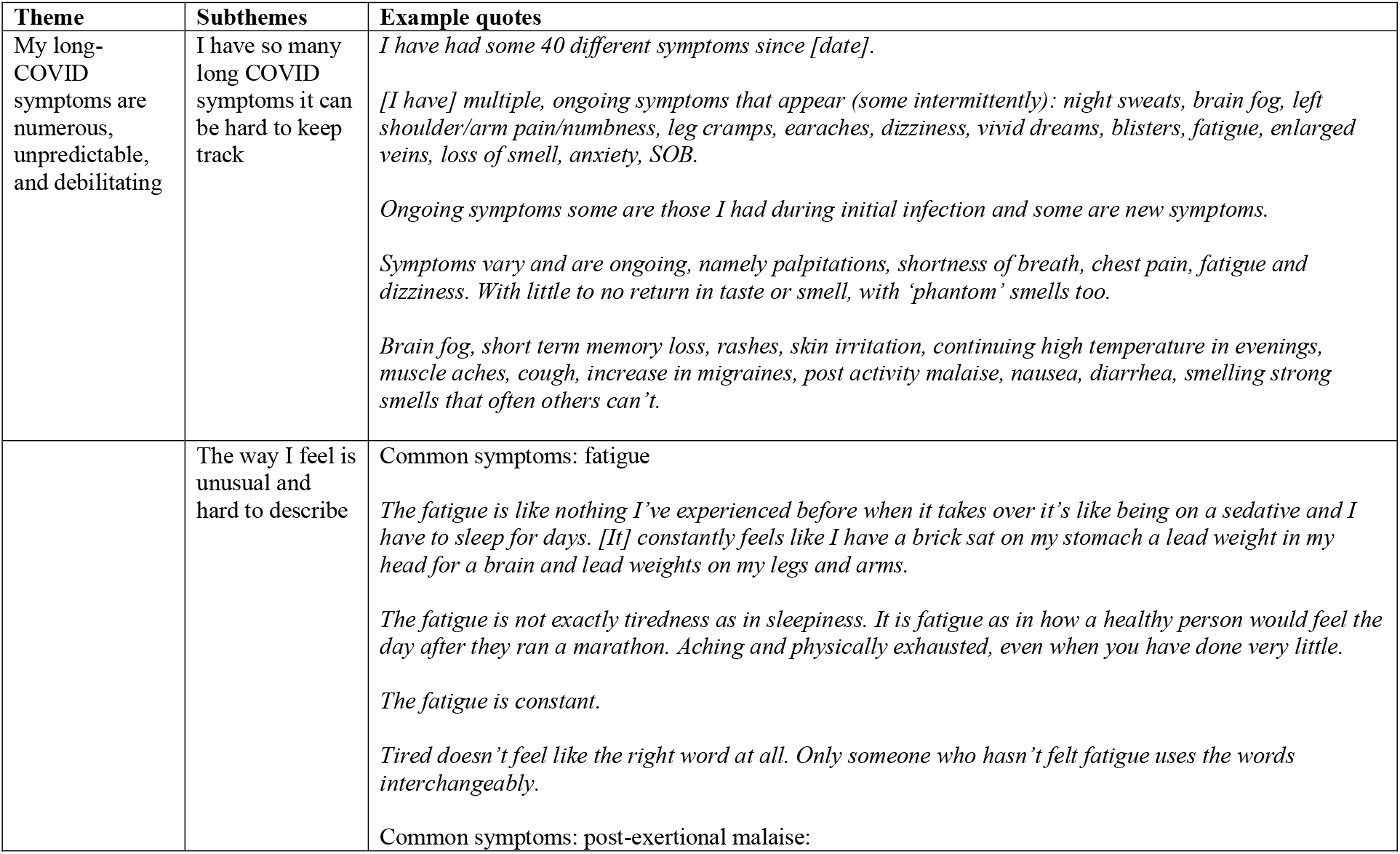
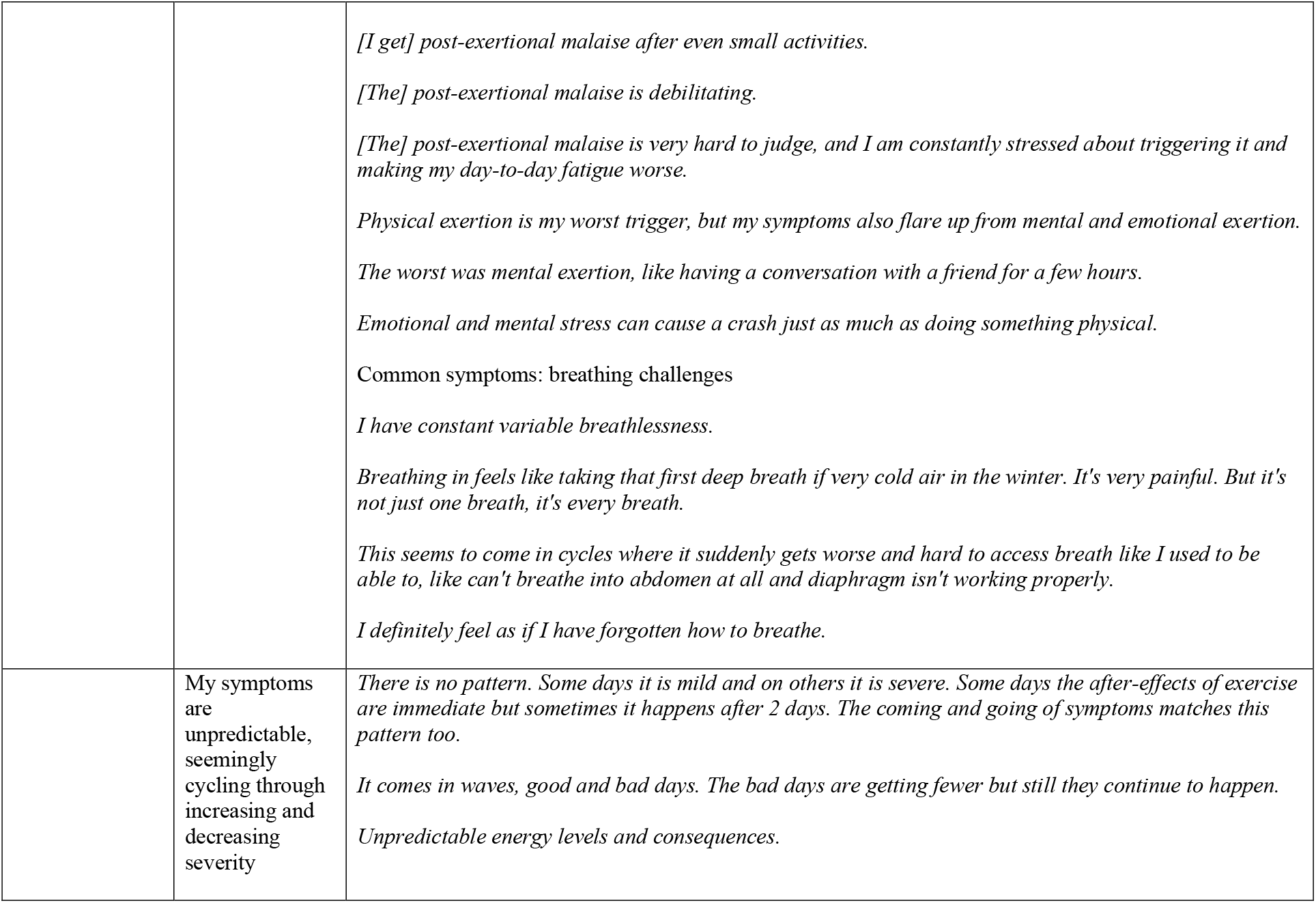
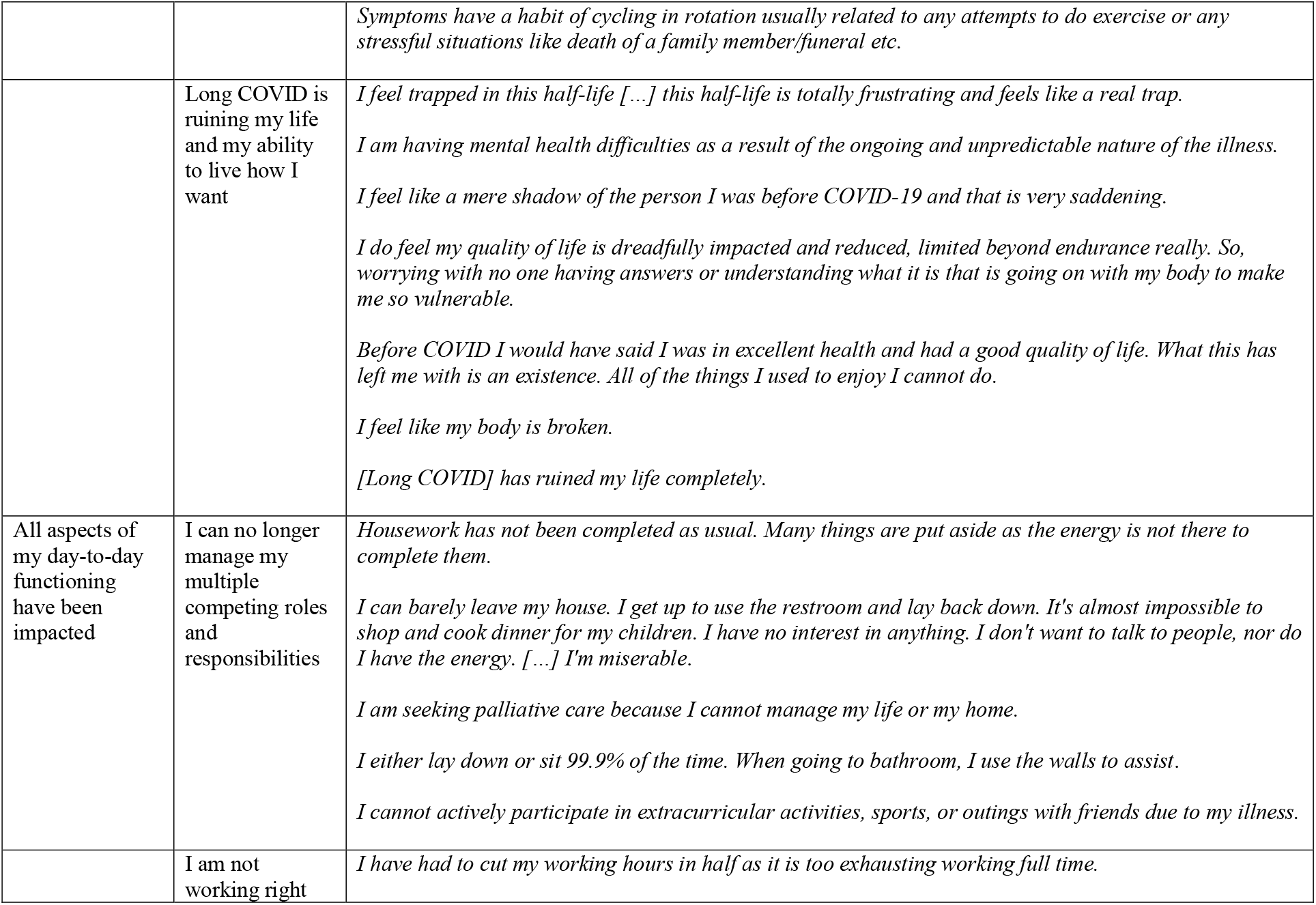
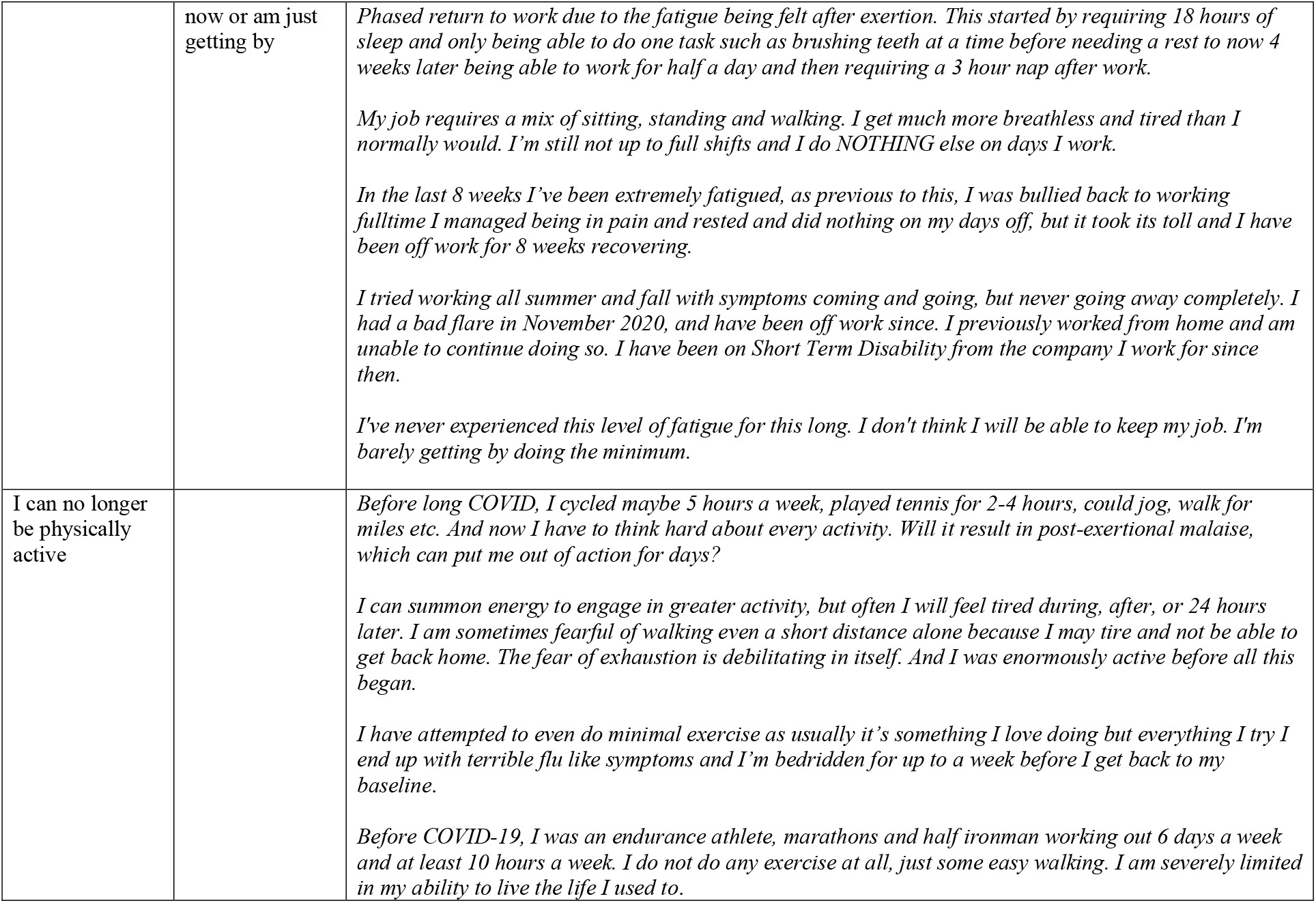
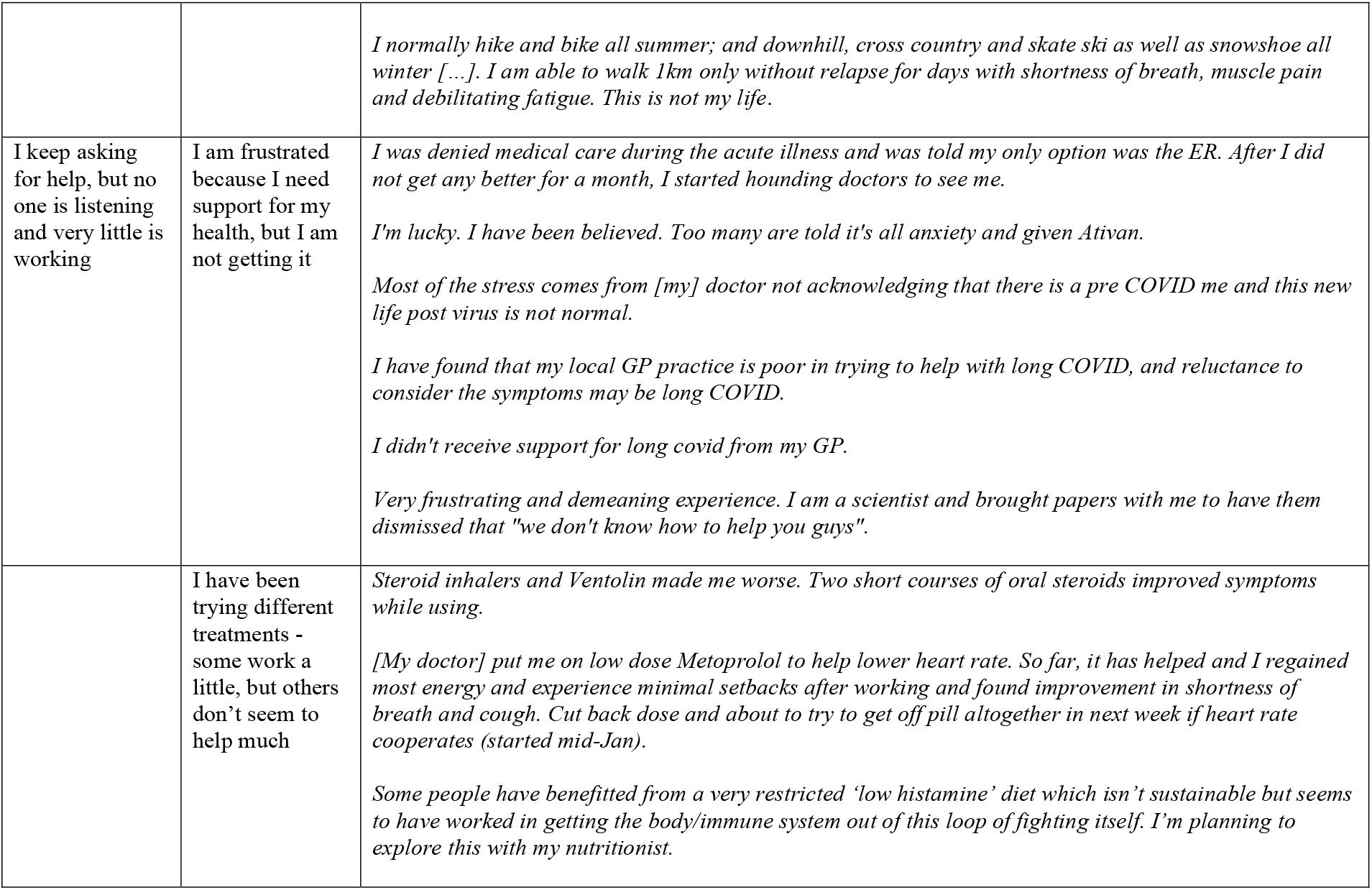
Additional illustrative quotations for themes and subthemes capturing participants’ experiences with long COVID and commonly reported symptoms.

#### 1. My Long-COVID Symptoms are Numerous, Hard to Describe, and Debilitating

This theme captures when participants described the unpredictable and debilitating nature of their illness and the ways in which their symptoms were distressing, multiple, and varied.

##### I have so many long COVID symptoms it can be hard to keep track

The quantity of symptoms participants had or were currently experiencing was described as overwhelming and unmanageable. This was highlighted by one participant who shared: “*I have had many, many more types of symptoms after the acute period, hundreds*”. While some participants attempted to list each of their symptoms, many indicated it was far too many to note. For example, one participant wrote, “*Neurological, short term memory loss, loss concentration, unable to retain information when reading, difficulty with math, dizziness, blurred vision, severe tinnitus with hearing loss, bilateral ulnar neuropathy, sleep apnea with hypoxia, abdominal pain and cramping, diarrhea, temperature intolerance heat and cold, toenails falling off. Fatigue is horrible. Daily constant headache. Lots more can’t remember*”, while another shared: “*There are too many symptoms to mention*.”

##### The way I feel is unusual and hard to describe

In response to open-ended items asking about common long COVID symptoms, including fatigue, post-exertional malaise, and breathing, participants shared that they were fatigued most, if not all, of the time. Participants explained how their fatigue was so much more than being tired, it was described as unrelenting and unlike anything else they had ever experienced. This was captured by a participant who wrote: “*I describe COVID and long COVID fatigue as crushing. So many days these last 8 months I’ve gotten out of bed only to be hit with a crushing wave of fatigue immediately that forces me to go back to bed*”. Further, participants shared that post-exertional malaise was experienced often, and was seemingly triggered by a wide range of activities, ranging from exercise (“*I feel ill and my original [COVID-19] symptoms return after exercise*”) to cognitive tasks (“*I do get fatigue and worsened symptoms including sore throat, stuffy nose, headache, aching, fatigue, dizziness after doing cognitive tasks for a long period, such as writing and reading*”). Finally, participants shared their variable breathing challenges. For example, one participant shared: “*I feel constantly breathless […]. I often have an expiratory wheeze. I feel that I cannot fully exhale, inhalation feels pretty fully but unfulfilling*,” and another shared: “*I get breathless talking to people, moving around or exerting myself (e*.*g*., *washing, dressing). I am ok when I am resting quietly*”.

##### My symptoms are unpredictable, seemingly cycling through increasing and decreasing severity

Participants offered their perspective on the nature of long COVID when they recounted navigating relapsing and remitting symptoms, which for many, were completely unpredictable. This sentiment was captured by one participant who wrote: “*It’s unpredictable. One day I can be energetic and the next, exhausted*”. This was elaborated on by another participant who wrote: “*Most [symptoms] I continue to have. Most are debilitating, painful, or bizarre. They are often cyclical fading and getting worse again while others take their place doing the same*”.

##### Long COVID is ruining my life and my ability to live how I want

The debilitating impact of long COVID on participants’ lives, in general, was made apparent and discussed as a major concern. Participants expressed a deep sense of loss over their changed health and reduced quality of life, which is illustrated by one participant who shared: “*I was a happy, healthy, fit person prior to developing this virus. My life has been turned upside down and I have no idea if I will ever get better*”. Many expressed feeling trapped or stuck within a disabled body that they no longer recognize nor understand, which was experienced as frustrating and scary. One participant wrote: “*I don’t know this person I have become since COVID-19*,” and another shared: “*I now consider myself disabled in many ways*.”

#### 2. All Aspects of my Day-To-Day Functioning Have Been Impacted

This theme captures when participants mentioned noticing marked changes in their functional abilities and capacity to maintain their responsibilities and roles. Consequently, participants described the many ways they modified their day or accepted being unable to complete necessary activities of daily living.

##### I can no longer manage my multiple competing roles and responsibilities

Participants indicated they could no longer take care of their home, families, and in some cases, themselves as a result of their symptoms. For many participants, their current functional capacity stood in stark contrast to their life before COVID-19. One participant illustrated this when they shared: “*My life is completely different than 1 year ago. I used to work full time, do wedding hair part-time, and had friends and many hobbies. Now I am unable to work, can barely shower without resting after, and sometimes I can’t even get my own groceries or cook for myself*”. Participants also described that despite social opportunities being limited due to physical distancing restrictions, they are still unable to manage engaging in, or thinking about, being social. One participant wrote: “*I cannot enjoy any time with my family and friends anymore, and even if I could, I get so tired I would have a window of about 20 minutes*”. Being unable to manage their day-to-day life was described by participants as extremely upsetting and distressing, which was captured by a participant who wrote: “*To avoid days of relapse, I literally do almost nothing. This is not who I am, and not what my primary relationship was based on. [It is] very depressing at times*”.

##### I am not working right now or am just getting by

Most participants within this sample wrote that they were not working or had greatly reduced the hours within their workweek to help manage their long COVID symptoms. For those participants who were not currently working, concerns over lost income and/or being dismissed were prevalent, which was captured by a participant who shared: “*I fear the loss of income as I’ve been off work for 3 months and am now subject to occupational health adviser reports to my employer with the risk of dismissal due to ill health*”. For those participants who were working, many described phased returns to work, needing modified schedules, working from bed or the couch, or being completely depleted after work and on their days off. One participant wrote: “*I only work 4 days per week, but post-exertional malaise affects me all the time when I am no working, which is affecting quality of life. My home life and jobs [i*.*e*., *chores] are left undone and spiraling out of control*”.

#### 3. I Can no Longer be Physically Active

This theme captures when participants wrote about changing their physical activity behaviour as a result of post-exertional malaise or symptoms during physical activity and exercise. Participants expressed a sense of loss over their pre-COVID physical activity and exercise behaviour: “*To go from always moving, playing with children, walking everywhere, swimming, spinning, golf etc. to sitting down for 16 hours a day is devastating*”. Some participants expressed deep-seated fears of engaging in more physical activity in case it worsened their symptoms and caused further setbacks, which was underlined by a sense of deteriorating trust in their own bodies. This was captured by a participant who wrote: “*I tried to go for a 20-minute walk round the block, timed myself so not to do too much, it took me three days to recover enough to move about my small flat comfortably after that, so I got nervous about going out again. Now I just stay inside, scared that even the smallest amount of exercise is going to push me back into being in bed, immobile again. It is terrifying to have been physically active before and now finding myself so incapable of what was totally not anything I would have thought about before*”.

#### 4. I Keep Asking for Help, but no one is Listening, and Very Little is Working

This theme captures when participants described the support (or lack thereof) they received from the healthcare system. Participants expressed a desire to be heard and taken seriously and to be helped. Further, participants described what they had done/tried to minimize symptoms, whether under guidance from their healthcare team or alternative sources.

##### I am frustrated because I need support for my health, but I am not getting it

Participants recounted that they were ignored or dismissed by their primary healthcare providers, which left many feeling defeated, helpless, invisible, or frustrated. This was captured by a participant who wrote: “*For the first 9 months of long COVID, the fact that nobody knew anything about long COVID, how to treat it and whether it would be chronic increased a sense of hopelessness which definitely affected my mental health and quality of life*”. However, participants described still needing care, and so many have continued to advocate for themselves, despite their symptoms. Participants used the space within the open-ended items in this sub-study to ask, and in some cases, beg for help. For example, one participant wrote: “*I have been trying to get help, but when every test comes back normal, I have been told there is nothing to be done. With so many of us suffering someone has to help us. Please help us I beg you*”.

##### I have been trying different treatments - some work a little, but others don’t seem to help much

Participants described what they have done (or are doing) to minimize their symptoms. Unfortunately, many described their symptoms persisting regardless of their attempts to find treatment(s). One participant elucidated this when they wrote: “*I have been taking 10 mg of cetirizine and 1,000 mg of Quercetin for 2 weeks now (self-prescribed) and have noticed a difference in the fatigue, brain fog and lung congestion (though all still remain)*”. While some participants were of the opinion that pharmacological (e.g., medication) and non-pharmacological options (e.g., diet, physiotherapy) could offer minor relief from symptoms, for others, no treatment(s) had yet offered a reprieve. This was captured by a participant who wrote: “*Traditional medicine has been of no help. Relief to some extent from breathing physio, elimination of alcohol from diet, and Symbicort (though no asthma)*” and another who shared: “*I have been doing breathing exercises daily for 8 weeks but I can’t my peak flow above 300. [I] was on a rescue inhaler only for years. Since COVID I went up to three inhalers. Then went for respiratory testing. Now on Symbicort. 8 puffs/day for four weeks now and very little improvement*”.

## Discussion

The purpose of this sub-study was to better understand and explore individuals’ experiences with long COVID and commonly reported symptoms using qualitative data collected from open-ended survey items contained within a larger survey [8]. Findings support and extend prior research and underscore the complex, highly variable, and distressing nature of long COVID across a large number of participants from predominantly developed countries. Results also align with literature capturing the vexing experience of navigating an ‘invisible’ and poorly understood illness that disrupts nearly all aspects of daily living [24–27]. These findings contribute to a growing evidence base calling for continued efforts to better understand and improve care for individuals with long COVID.

Participants in this sample described their ‘new’ long COVID body as being discordant with their physical, occupational, and social obligations. This dissonance manifested as feelings of deep loss, sadness, and frustration. Mourning over one’s pre-illness identity and ability to participate in roles, responsibilities, and leisure pursuits has been reported previously with individuals affected by long COVID [25] and other chronic conditions, including fibromyalgia and myalgic encephalomyelitis/chronic fatigue syndrome (ME/CFS) [35–38]. Beyond sharing sentiments of loss, participants in this study also offered insights into how they have altered their day-to-day life to account for their reduced capacities and possible continued deterioration. For example, some participants described sacrificing their social activities to keep up at work; others had to leave work altogether or were fearful of termination; whereas others chose to engage in one task per day or to take breaks after every activity. These findings align with the recent conceptualization of long COVID as characterized by health-related challenges or episodic disability [39,40]. Looking ahead, exploring if and how individuals adapt to their post-illness body will be necessary. For example, post-exertional malaise can greatly limit the ability to engage in activities that could otherwise help prevent low mood and feelings of isolation [41]. When combined with severe functional limitations and the burden of educating others, there can be serious adverse impacts on mental health [41,42]. A deeper exploration of common symptoms, including post-exertional malaise, and their physical and psychological consequences, could disentangle the effects of long COVID from the secondary impacts on mental health - ultimately informing interventions and supportive care strategies.

There were multiple and varied ways participants described self-managing their condition, from consuming medical information and advocating for themselves to adhering to treatments that do not always offer relief. Participants also shared the additional challenge they faced in self-managing their condition within a system that challenged or contested their diagnosis. This sentiment of navigating one’s condition amidst doubts about authenticity has been reported in individuals with ME/CFS [43–47]. ME/CFS can occur after an infectious illness such as mononucleosis [48], but it remains a highly stigmatized health condition where patients are vulnerable to epistemic injustice in healthcare encounters [45]. The feelings of dismissal from healthcare providers expressed by participants in this sample, the conceptual ambiguity of both ME/CFS and long COVID, and the nature of symptoms which are numerous and hard to describe, may result in low credibility being given to the patient testimonial [45]. The recent recognition of long COVID as a real outcome of COVID-19 infection (for example, through being given a clinical case definition by the World Health Organization [32]) may help further legitimize the illness. Considering that long COVID is likely to lead to ME/CFS for some patients [49], an important next step is educating primary healthcare providers, who are the front line for patients, to recognize and validate their patients. Beyond this, equipping healthcare providers with evidence-based guidelines and referral pathways is necessary to better support a growing patient population as they adapt to long COVID.

However, educating primary healthcare providers with evidence-based guidelines and referral pathways may not be enough. There is a critical need to identify appropriate interventions and treatments, which may be best accomplished by including patients as partners in the development and evaluation of potential interventions and treatments. Anecdotally (and in the authors’ clinical experience), some individuals with long COVID are responding well to self-management, including activity pacing over extended time periods. Given that typical rehabilitation timeframes are often 2-3 months, and existing rehabilitation approaches (such as pulmonary rehabilitation) are unlikely to be suitable for the majority of people with long COVID because there are distinct clinical presentations [50], novel approaches may be necessary. For example, multidisciplinary, tailored interventions that leverage peer support may be most effective and should adhere to the recommended quality standards of being evidence-based, accessible and of minimal burden [26,27]. Occupational therapists, physiotherapists, and other rehabilitation specialists who have expertise and experience working with people who have been discredited in the past (e.g., fibromyalgia, ME/CFS or other chronic conditions resulting in drastically changed capacity or disability) may be best placed to work with patients to design appropriate and effective interventions and treatments. Furthermore, appropriate training of all professionals working with people with long COVID may enhance continuity of care and offer prolonged support for this cohort (as opposed to the often disconnected care pathways). Engaging with patients and learning from their experiences represents a valuable first step towards improving training and care.

### Considerations

Although this study contributes to and extends prior research, there are a number of considerations that must be taken into account. First, as described in the manuscript describing the larger study [8], studies using online questionnaires are subject to selection bias. Thus, the extent to which the sample described herein is representative of the population of people living with long COVID is unknown. Indeed, this sample primarily self-identified as White women and as residing in North America or Europe. Gaining insights from those with different backgrounds and residing in different countries is therefore necessary. Further, finding ways to capture perspectives from historically marginalized and underrepresented groups is critical to fully understand long COVID and its potential range of impacts. Second, this sub-study collected and analyzed open-ended responses to items presented after closed-ended questionnaires (assessing specific outcomes) or blocks of questions. It is, therefore, possible that the open-ended item stems were leading, prompting participants to respond using a specific language. A number of efforts were made to minimize this. For example, the item stems prompted participants to reflect on their broader experiences and use the space if they had additional insights to share and/or to clarify their responses to closed-ended items. As well, participants were reminded throughout that their response to open-ended items was completely optional. Third, online open-ended items do not allow the authors to probe to clarify meaning or gain deeper insights. Nevertheless, the authors felt that the benefits of using online open-ended items (i.e., collating a large number of firsthand accounts, low participant burden, anonymity) outweighed these considerations and aligned with the pragmatic approach adopted wherein a balance was sought between advancing conceptualizations of long COVID, and sharing evidence to support this population in real-time.

## Conclusion

Given the uncertain nature of long COVID, exploring individuals’ lived experience is paramount. Findings reiterate that long COVID is complex and distressing for those affected. The varied relapsing-remitting symptoms, unknown prognosis and deep sense of loss over one’s prior identity suggest interventions are needed to support this population. Further, results underscore the challenges individuals affected by long COVID face when advocating for themselves and adapting to their illness during the pandemic and amidst healthcare systems that are understaffed, at times disbelieving or unarmed (as of yet) with comprehensive treatment guidelines. More research is needed to identify and address the pathophysiology, capture the consequences of long COVID, implement strategies to support those affected, and ultimately better help this cohort navigate the process of adapting to long COVID.

## Data Availability

Qualitative data are summarized in the manuscript. De-identified quantitative data for the wider study are available online at: https://osf.io/dxu63/.

## CRediT author statement

Amanda Wurz: Conceptualization, Methodology, Software, Formal analysis, Data curation, Writing - original draft. Jessica DeMars: Conceptualization, Writing - review and editing.

Kelli Franklin: Patient partner (provided guidance based on the lived experience of long COVID), Validation, Writing - review and editing.

S. Nicole Culos-Reed: Writing - review and editing.

James G. Wrightson: Conceptualization, Writing - review and editing.

Rosie Twomey: Conceptualization, Methodology, Software, Validation, Data curation, Writing - review and editing, Project administration.

## Acknowledgements

The authors wish to sincerely thank the people living with long COVID who took part in this study for sharing their experiences.

## References

1. Zaim S, Chong JH, Sankaranarayanan V, Harky A. COVID-19 and Multiorgan Response. Curr Probl Cardiol. 2020;45: 100618. doi:10.1016/j.cpcardiol.2020.100618

2. Wiersinga WJ, Rhodes A, Cheng AC, Peacock SJ, Prescott HC. Pathophysiology, Transmission, Diagnosis, and Treatment of Coronavirus Disease 2019 (COVID-19): A Review. JAMA. 2020;324: 782–793. doi:10.1001/jama.2020.12839

3. Prevalence of ongoing symptoms following coronavirus (COVID-19) infection in the UK - Office for National Statistics. [cited 23 May 2021]. Available: https://www.ons.gov.uk/peoplepopulationandcommunity/healthandsocialcare/conditionsanddiseases/bulletins/prevalenceofongoingsymptomsfollowingcoronaviruscovid19infectionintheuk/1april2021

4. Callard F, Perego E. How and why patients made Long Covid. Social Science & Medicine. 2021;268: 113426. doi:10.1016/j.socscimed.2020.113426

5. Dennis A, Wamil M, Alberts J, Oben J, Cuthbertson DJ, Wootton D, et al. Multiorgan impairment in low-risk individuals with post-COVID-19 syndrome: a prospective, community-based study. BMJ Open. 2021;11: e048391. doi:10.1136/bmjopen-2020-048391

6. NICE. COVID-19 rapid guideline: managing the long-term effects of COVID-19. NICE; 18 Dec 2020 [cited 19 May 2021]. Available: https://www.nice.org.uk/guidance/ng188/chapter/common-symptoms-of-ongoing-symptomatic-covid-19-and-post-covid-19-syndrome#common-symptoms-of-ongoing-symptomatic-covid-19-and-post-covid-19-syndrome

7. Davis HE, Assaf GS, McCorkell L, Wei H, Low RJ, Re’em Y, et al. Characterizing long COVID in an International cohort: 7 months of symptoms and their impact. medRxiv. 2020; 2020.12.24.20248802. doi:10.1101/2020.12.24.20248802

8. Twomey R, DeMars J, Franklin K, Culos-Reed SN, Weatherald J, Wrightson JG. Chronic fatigue and post-exertional malaise in people living with long COVID. Physical Therapy (In Press). 2021; 2021.06.11.21258564. doi:10.1101/2021.06.11.21258564

9. Gorna R, MacDermott N, Rayner C, O’Hara M, Evans S, Agyen L, et al. Long COVID guidelines need to reflect lived experience. The Lancet. 2021;397: 455–457. doi:10.1016/S0140-6736(20)32705-7

10. Brown DA, O’Brien KK, Josh J, Nixon SA, Hanass-Hancock J, Galantino M, et al. Six Lessons for COVID-19 rehabilitation from HIV rehabilitation. Physical Therapy. 2020;100: 1906–1909. doi:10.1093/ptj/pzaa142

11. Taylor AK, Kingstone T, Briggs TA, O’Donnell CA, Atherton H, Blane DN, et al. “Reluctant pioneer”: A qualitative study of doctors’ experiences as patients with long COVID. Health Expectations. [cited 16 May 2021]. doi:10.1111/hex.13223

12. Alwan NA. Surveillance is underestimating the burden of the COVID-19 pandemic. The Lancet. 2020;396: e24. doi:10.1016/S0140-6736(20)31823-7

13. Alwan NA. The teachings of Long COVID. Commun Med. 2021;1: 1–3. doi:10.1038/s43856-021-00016-0

14. Davis HE, Assaf GS, McCorkell L, Wei H, Low RJ, Re’em Y, et al. Characterizing long COVID in an international cohort: 7 months of symptoms and their impact. EClinicalMedicine. 2021;0. doi:10.1016/j.eclinm.2021.101019

15. World Physiotherapy. World Physiotherapy Response to COVID-19 Briefing Paper 9. Safe rehabilitation approaches for people living with Long COVID: physical activity and exercise. London, UK: World Physiotherapy; 2021.

16. Nalbandian A, Sehgal K, Gupta A, Madhavan MV, McGroder C, Stevens JS, et al. Post-acute COVID-19 syndrome. Nature Medicine. 2021;27: 601–615. doi:10.1038/s41591-021-01283-z

17. Tabacof L, Tosto-Mancuso J, Wood J, Cortes M, Kontorovich A, McCarthy D, et al. Post-acute COVID-19 syndrome negatively impacts health and wellbeing despite less severe acute infection. medRxiv. 2020; 2020.11.04.20226126. doi:10.1101/2020.11.04.20226126

18. Huang C, Huang L, Wang Y, Li X, Ren L, Gu X, et al. 6-month consequences of COVID-19 in patients discharged from hospital: a cohort study. The Lancet. 2021;397: 220–232. doi:10.1016/S0140-6736(20)32656-8

19. Li J, Xia W, Zhan C, Liu S, Yin Z, Wang J, et al. Effectiveness of a telerehabilitation program for COVID-19 survivors (TERECO) on exercise capacity, pulmonary function, lower limb muscle strength, and quality of life: a randomized controlled trial. medRxiv. 2021; 2021.03.08.21253007. doi:10.1101/2021.03.08.21253007

20. Group P-CC, Evans RA, McAuley H, Harrison EM, Shikotra A, Singapuri A, et al. Physical, cognitive and mental health impacts of COVID-19 following hospitalisation – a multi-centre prospective cohort study. medRxiv. 2021; 2021.03.22.21254057. doi:10.1101/2021.03.22.21254057

21. Sykes DL, Holdsworth L, Jawad N, Gunasekera P, Morice AH, Crooks MG. Post-COVID-19 Symptom Burden: What is Long-COVID and How Should We Manage It? Lung. 2021;199: 113–119. doi:10.1007/s00408-021-00423-z

22. Alwan NA, Johnson L. Defining long COVID: Going back to the start. Med (N Y). 2021;2: 501–504. doi:10.1016/j.medj.2021.03.003

23. Rando HM, Bennett TD, Byrd JB, Bramante C, Callahan TJ, Chute CG, et al. Challenges in defining Long COVID: Striking differences across literature, Electronic Health Records, and patient-reported information. medRxiv. 2021; 2021.03.20.21253896. doi:10.1101/2021.03.20.21253896

24. Kingstone T, Taylor AK, O’Donnell CA, Atherton H, Blane DN, Chew-Graham CA. Finding the “right” GP: a qualitative study of the experiences of people with long-COVID. BJGP Open. 2020;4. doi:10.3399/bjgpopen20X101143

25. Humphreys H, Kilby L, Kudiersky N, Copeland R. Long COVID and the role of physical activity: a qualitative study. BMJ Open. 2021;11: e047632. doi:10.1136/bmjopen-2020-047632

26. Ladds E, Rushforth A, Wieringa S, Taylor S, Rayner C, Husain L, et al. Developing services for long COVID: lessons from a study of wounded healers. Clin Med (Lond). 2021;21: 59–65. doi:10.7861/clinmed.2020-0962

27. Ladds E, Rushforth A, Wieringa S, Taylor S, Rayner C, Husain L, et al. Persistent symptoms after Covid-19: qualitative study of 114 “long Covid” patients and draft quality principles for services. BMC Health Services Research. 2020;20: 1144. doi:10.1186/s12913-020-06001-y

28. Braun V, Clarke V. Reflecting on reflexive thematic analysis. Qualitative Research in Sport, Exercise and Health. 2019;11: 589–597. doi:10.1080/2159676X.2019.1628806

29. Braun V, Clarke V, Boulton E, Davey L, McEvoy C. The online survey as a qualitative research tool. International Journal of Social Research Methodology. 2020;0: 1–14. doi:10.1080/13645579.2020.1805550

30. Kelly LM, Cordeiro M. Three principles of pragmatism for research on organizational processes. Methodological Innovations. 2020;13: 2059799120937242. doi:10.1177/2059799120937242

31. Creswell JW & Poth CN 2013. Philosophical assumptions and interpretive frameworks (chapter 2). In Qualitative Inquiry & Research Design 4th ed. Thousand Oaks, California; Sag.

32. World Health Oganization (2021). A clinical case definition of post COVID-19 condition by a Delphi consensus. Accessed: Oct 6, 2021. Available: https://www.who.int/publications/i/item/WHO-2019-nCoV-Post_COVID-19_condition-Clinical_case_definition-2021.1.

33. The jamovi project (2021). jamovi (Version 1.6) [Computer Software]. Retrieved from https://www.jamovi.org.

34. Yardley L. Dilemmas in qualitative health research. Psychology & Health. 2000;15: 215–228. doi:10.1080/08870440008400302

35. Ashe SC, Furness PJ, Taylor SJ, Haywood-Small S, Lawson K. A qualitative exploration of the experiences of living with and being treated for fibromyalgia. Health Psychol Open. 2017;4: 2055102917724336. doi:10.1177/2055102917724336

36. Alameda Cuesta A, Pazos Garciandía Á, Oter Quintana C, Losa Iglesias ME. Fibromyalgia, Chronic Fatigue Syndrome, and Multiple Chemical Sensitivity: Illness Experiences. Clin Nurs Res. 2021;30: 32–41. doi:10.1177/1054773819838679

37. Bartlett C, Hughes JL, Miller L. Living with myalgic encephalomyelitis/chronic fatigue syndrome: Experiences of occupational disruption for adults in Australia. British Journal of Occupational Therapy. 2021; 03080226211020656. doi:10.1177/03080226211020656

38. Sandhu RK, Sundby M, Ørneborg S, Nielsen SS, Christensen JR, Larsen AE. Lived experiences in daily life with myalgic encephalomyelitis. British Journal of Occupational Therapy. 2021;84: 658–667. doi:10.1177/0308022620966254

39. Brown DA, O’Brien KK. Conceptualising Long COVID as an episodic health condition. BMJ Global Health. 2021;6: e007004. doi:10.1136/bmjgh-2021-007004

40. O’Brien KK, Bayoumi AM, Strike C, Young NL, Davis AM. Exploring disability from the perspective of adults living with HIV/AIDS: development of a conceptual framework. Health Qual Life Outcomes. 2008;6: 76. doi:10.1186/1477-7525-6-76

41. Chu L, Elliott M, Stein E, Jason LA. Identifying and Managing Suicidality in Myalgic Encephalomyelitis/Chronic Fatigue Syndrome. Healthcare (Basel). 2021;9: 629. doi:10.3390/healthcare9060629

42. Devendorf AR, McManimen SL, Jason LA. Suicidal ideation in non-depressed individuals: The effects of a chronic, misunderstood illness. J Health Psychol. 2020;25: 2106–2117. doi:10.1177/1359105318785450

43. McManimen S, McClellan D, Stoothoff J, Gleason K, Jason LA. Dismissing chronic illness: A qualitative analysis of negative health care experiences. Health Care Women Int. 2019;40: 241–258. doi:10.1080/07399332.2018.1521811

44. McManimen SL, McClellan D, Stoothoff J, Jason LA. Effects of unsupportive social interactions, stigma, and symptoms on patients with myalgic encephalomyelitis and chronic fatigue syndrome. J Community Psychol. 2018;46: 959–971. doi:10.1002/jcop.21984

45. Blease C, Carel H, Geraghty K. Epistemic injustice in healthcare encounters: evidence from chronic fatigue syndrome. J Med Ethics. 2017;43: 549–557. doi:10.1136/medethics-2016-103691

46. Bayliss K, Goodall M, Chisholm A, Fordham B, Chew-Graham C, Riste L, et al. Overcoming the barriers to the diagnosis and management of chronic fatigue syndrome/ME in primary care: a meta synthesis of qualitative studies. BMC Fam Pract. 2014;15: 44. doi:10.1186/1471-2296-15-44

47. Bayliss K, Riste L, Fisher L, Wearden A, Peters S, Lovell K, et al. Diagnosis and management of chronic fatigue syndrome/myalgic encephalitis in black and minority ethnic people: a qualitative study. Prim Health Care Res Dev. 2014;15: 143–155. doi:10.1017/S1463423613000145

48. Jason LA, Yoo S, Bhatia S. Patient perceptions of infectious illnesses preceding Myalgic Encephalomyelitis/Chronic Fatigue Syndrome. Chronic Illn. 2021; 17423953211043106. doi:10.1177/17423953211043106

49. Komaroff AL, Bateman L. Will COVID-19 Lead to Myalgic Encephalomyelitis/Chronic Fatigue Syndrome? Front Med. 2021;7. doi:10.3389/fmed.2020.606824

50. Whitaker M, Elliott J, Chadeau-Hyam M, Riley S, Darzi A, Cooke G, et al. Persistent symptoms following SARS-CoV-2 infection in a random community sample of 508,707 people. 2021 Jul p. 2021.06.28.21259452. doi:10.1101/2021.06.28.21259452

